# *NEUROD1* and *PDX1* are low penetrance causes of MODY while rare variants in *APPL1* and *WFS1* are not associated with MODY

**DOI:** 10.1101/2025.05.07.25327066

**Authors:** Aparajita Sriram, Matthew N. Wakeling, Andrew T. Hattersley, Michael N. Weedon, Kevin Colclough, Thomas W. Laver, Kashyap A. Patel

## Abstract

An accurate genetic diagnosis of Maturity-Onset Diabetes of the Young (MODY) is critical for personalised treatment. To avoid misdiagnosis, only genes with strong evidence of causality must be tested. Heterozygous variants in *NEUROD1*, *PDX1*, *APPL1*, and *WFS1* have been implicated in MODY, but strong genetic evidence supporting causality is lacking. We therefore assessed their existing genetic evidence and performed the gene-level burden tests in a large MODY cohort, alongside two established MODY genes as positive controls (*HNF1A*-high penetrance, *RFX6* -low penetrance).

The first reported MODY-associated variants in *NEUROD1*, *PDX1*, *APPL1*, and *WFS1* were <1:20,000 frequency. Based on the small number of large published pedigrees (n<3), MODY-associated variants showed only modest co-segregation in these genes. Crucially, ultra-rare (MAF<1:10,000) protein-truncating and predicted-damaging missense variants in *APPL1* and *WFS1* were not enriched in a MODY cohort (n=2,571) compared to population controls (n=155,501; all p>0.06). In contrast, variants in *NEUROD1* and *PDX1* were enriched, albeit at levels comparable to *RFX6*. Multiple sensitivity analyses corroborated these findings. In summary, rare heterozygous variants in *NEUROD1* and *PDX1* are low-penetrance causes of MODY, while those in *APPL1* and *WFS1* lack robust genetic evidence for causality and should not be included in MODY testing panels.

**Article highlights:**

a. **Why did we undertake this study?** Testing genes with limited evidence of causality risks misdiagnosis of MODY.
b. **What is the specific question(s) we wanted to answer?** Do rare variants in *PDX1*, *NEUROD1*, *APPL1*, and *WFS1* cause MODY?
c. **What did we find?** Rare damaging variants in *APPL1* and *WFS1* are not enriched in a MODY cohort, but those in *PDX1* and *NEUROD1* are, at a level similar to low-penetrance MODY genes.
d. **What are the implications of our findings?** *PDX1* and *NEUROD1* should be included in MODY gene panels, while heterozygous *APPL1* and *WFS1* variants should not be reported as causes of MODY.

## Introduction

An accurate genetic diagnosis of Maturity Onset Diabetes of the Young (MODY) is vital for patients but requires knowledge of the causal genes to prevent misdiagnosis. MODY is the most common form of monogenic diabetes, responsible for ∼3% of all diabetes diagnosed under 30 years old (1). MODY is an autosomal dominant, genetically heterogeneous disease with 16 genes implicated to date (2). A genetic diagnosis optimises treatment, for example, individuals with HNF1A-MODY respond well to sulfonylurea tablets (3). Currently, genetic testing is primarily conducted using a multi-gene panel approach, where all relevant genes are analysed in a single assay. However, testing genes with limited evidence of causality increases the risk of misdiagnosis and inappropriate clinical management.

Re-evaluating genetic evidence can identify MODY genes without sufficient evidence for causality. We recently demonstrated that variants in *BLK*, *KLF11,* and *PAX4* reported to cause MODY are common in the population, have limited co-segregation, and lack enrichment in MODY cases compared to controls (4). Our approach leveraged large population databases such as the UK Biobank (5) and gnomAD (6) which were not available when these genes were published. This evidence contributed to the ClinGen international consensus panel (7) reclassifying the gene-disease relationships as refuted.

*NEUROD1*, *PDX1*, *APPL1* and *WFS1* have limited evidence of causality and need re-evaluation. Recently ClinGen identified *NEUROD1, PDX1* and *APPL1* as MODY genes where further genetic evidence is needed to either support or refute their pathogenicity (8). These genes were either published before large-scale population data was available and/or lacked ultra-rare variant enrichment analysis in MODY cases against population controls. This is also the case for heterozygous variants in *WFS1*. *WFS1* is a well-established cause of monogenic diabetes. However, the causal variants are either recessive loss-of-function causing Wolfram syndrome or specific *de novo* missense variants causing a syndrome of neonatal diabetes, deafness and cataracts (9). Dominant variants have been reported to cause adult-onset diabetes (10) but lack robust genetic evidence to support pathogenicity. Re-evaluating the evidence of causality in these genes is important to prevent misdiagnosis.

In this study, we used a large clinically suspected MODY cohort and population controls to assess the genetic evidence for *NEUROD1, PDX1, APPL1*, and *WFS1*.

## Research Design and Methods

### Study populations

#### MODY cohort

This cohort comprises 2,571 unrelated individuals of European ancestry with clinically suspected MODY from routine clinical practice, referred for genetic testing to the Exeter Genomics Laboratory (clinical features in Supplementary Table 1). The North Wales Ethics committee approved the study (no. 17/WA/0327). We obtained informed consent from probands or guardians.

#### UK Biobank

The UK Biobank is a population-based research initiative in the UK that recruited participants between the ages of 40 and 70. It has extensive phenotypic data along with comprehensive genetic information. For this study, we included individuals with genome sequencing data from the 200,000 genomes release of the UK Biobank. All participants provided informed consent, and the UK Biobank resource was approved by the UK Biobank Research Ethics Committee.

#### GnomAD

We used the Genome Aggregation Database (gnomAD) version 3.1.2 (6), which includes genome sequencing data from 76,156 individuals, as an alternative population control for a sensitivity analysis. GnomAD aggregates genetic data from various sequencing projects and makes it available to the wider scientific community.

### Genetic testing

#### MODY cohort

The probands underwent targeted next-generation sequencing for a panel of monogenic diabetes genes using a previously described method (11). We annotated variants using Alamut Batch 1.11 (Interactive Biosoftware, Rouen, France) against Genome Reference Consortium Human Build 37 (GRCh37) and lifted them over to Genome Reference Consortium Human Build 38 (GRCh38), to compare them with our controls. We examined the following RefSeq transcripts for our genes of interest: *APPL1* NM_012096.3, *HNF1A* NM_000545.8, *NEUROD1* NM_002500.5, *PDX1* NM_000209.4, *RFX6* NM_173560.4, and *WFS1* NM_006005.3.

#### UK Biobank

We included whole genome sequencing data from 155,501 unrelated individuals of European ancestry in the analysis. We focused on genome sequencing over exome sequencing data due to its well-described advantage of uniform coverage of coding exons to reduce the technical artefacts in our analysis (12) The sequencing methodology for UK Biobank is described in detail by Szustakowski *et al.* (13) and is available from https://biobank.ctsu.ox.ac.uk/showcase/label.cgi?id=170. Variant-calling was performed against genome build GRCh38. Variants were annotated using Alamut Batch 1.11 (Interactive Biosoftware, Rouen, France).

#### GnomAD

We included whole genome sequencing data from 34,029 Non-Finnish Europeans from gnomAD v3.1.2 to match with our MODY cohort. The gnomAD consortium conducted joint variant calling across samples using a standardized BWA-Picard-GATK pipeline. GnomAD performed quality control and analysis with the Hail open-source framework for scalable genetic analysis (6). Variants from gnomAD v3 were called against GRCh38. We annotated the gnomAD variants using Alamut Batch 1.11 (Interactive Biosoftware, Rouen, France).

#### Ultra-rare variant burden analysis

We performed stringent variant and sample-level quality control in all three cohorts to ensure that only high-quality variants were included in the analysis and to prevent technical artefacts from affecting our results. We focused on European ancestry across all the cohorts to prevent false inflation of statistics due to ancestral differences across the cohort. We have provided a detailed description of quality control steps in the Supplemental Methods. We performed ultra-rare variant (MAF <1:10,000) gene-level burden tests between the MODY cohort and UK Biobank controls and gnomAD v3.1.2 controls. We analyzed protein-truncating variants (PTVs), damaging missense variants defined using a REVEL score cutoff >0.7 (14) and synonymous variants in each gene. PTVs included stop-gain, splice site and frameshift variants. The primary analysis excluded PTVs expected to avoid nonsense mediated decay (NMD): those in the last exon and the last 50 base pairs of the penultimate exon. All PTVs in *NEUROD1* were included in the analysis because this is a single exon gene. We used the fisher’s exact test to assess the association. We also computed odds ratios (ORs) with 95% confidence intervals (CIs) to quantify effect sizes.

We used synonymous variants to assess test inflation and potential technical artefacts across cohorts. As positive controls in our analysis, we included *HNF1A* variants, representing a high-penetrance MODY gene, and *RFX6* variants, representing a low-penetrance MODY gene. To account for multiple testing, we applied a Bonferroni-corrected p-value threshold of 0.002 (0.05 / (6 genes × 3 groups)).

We performed power calculations to assess the detectable odds ratios at different minor allele frequencies. For variants with a MAF of 1 in 10,000, we had greater than 80% power to detect odds ratios as low as 9.8. At a MAF of 1 in 20,000, we maintained over 80% power to detect odds ratios of 14.5 or greater. For more common variants with a MAF of 1 in 5,000, we had sufficient power to detect odds ratios as low as 6.7.

#### Sensitivity Analysis for Burden Testing

We performed multiple sensitivity analyses. This included burden testing at both a stricter allele frequency threshold (MAF<1:20,000) and a more lenient threshold (MAF<1:5,000) to ensure that our findings were not dependent on the exact frequency cutoff chosen. Additionally, we conducted burden tests using gnomAD v3.1.2 controls as a sensitivity analysis, to test whether our results remained consistent with an independent control dataset. To further investigate potential region-specific effects, we performed burden tests focusing on functional domains within genes (Supplementary Methods). Finally, we conducted burden tests restricted to NMD escape regions to explore an alternative mechanism of disease.

### Re-evaluation of published data

#### Variant frequencies

We assessed the frequencies of the first published variants in our genes of interest. To check variant frequencies, we used gnomAD v2.1.1, which includes sequencing data from 141,456 individuals but excludes UK Biobank data to prevent overlap with our burden test analyses. The most common pathogenic MODY variant with at least 50% penetrance should appear no more than seven times in gnomAD v2.1.1 (frequency < 4.9 × 10⁻⁵) as per the framework from Whiffin *et al.* (15), using the MODY prevalence of 248 cases per million (16). However, it is important to note that variants in less common genes, like those analyzed in this study, which only contribute to a small proportion of MODY and have high variant heterogeneity, are expected to be much less frequent (17).

#### Co-segregation analysis

We reviewed all published pedigrees for putative MODY variants in our genes of interest to evaluate co-segregation, which measures how often a variant and disease are inherited together within a family. We followed ClinGen guidelines for this analysis and included only large pedigrees with four or more co-segregations (18). To avoid confounding the co-segregation results with common variants, we restricted our analysis to variants with a frequency below 1 in 1,500 in gnomAD v2.1.1. This exclusion criterion removed four pedigrees (19–22). We performed binomial tests to assess whether co-segregation (the proportion of family members with diabetes and the variant) and penetrance (the proportion of family members with the variant but without diabetes) were significantly different from the expected values of 50% and arbitrary threshold of 10%, respectively. The proportion of family members with diabetes and variant was calculated as the number of family members with both diabetes and variant divided by the total number of family members with diabetes and genotype information. The proportion of family members with the variant but without diabetes was calculated as the number of family members without diabetes with variant divided by the total number of family members without diabetes and genotype information who are at least 25 years of age.

## Results

### The first reported variants in NEUROD1, PDX1, APPL1 and WFS1 are rare in the population

Variants reported to cause MODY should be rarer than the disease itself. Therefore, we assessed the frequencies of the first published MODY variants in 141,456 individuals from gnomAD v2.1.1. We used *HNF1A* and *RFX6* as positive controls, representing highly penetrant and lower penetrant MODY genes, respectively. The first published variants in *NEUROD1* (23), *PDX1* (24), *APPL1* (*25*) and *WFS1* (10) are all rare in the population (allele frequency < 3x10) (Table 1). However, the first *APPL1* variants were published after the release of the first large population genetic database, ExAC (26), so would only have analyzed rare variants (25). The first *WFS1* variant shown to cause MODY was reported in a Finnish family before gnomAD but remains absent in 12,562 Finnish individuals in gnomAD (10). The *PDX1* variant was only seen in two individuals, and the NEUROD1 variant was present in six individuals but filtered out in GnomAD2.1.1. (23,24).

**Table 1:**
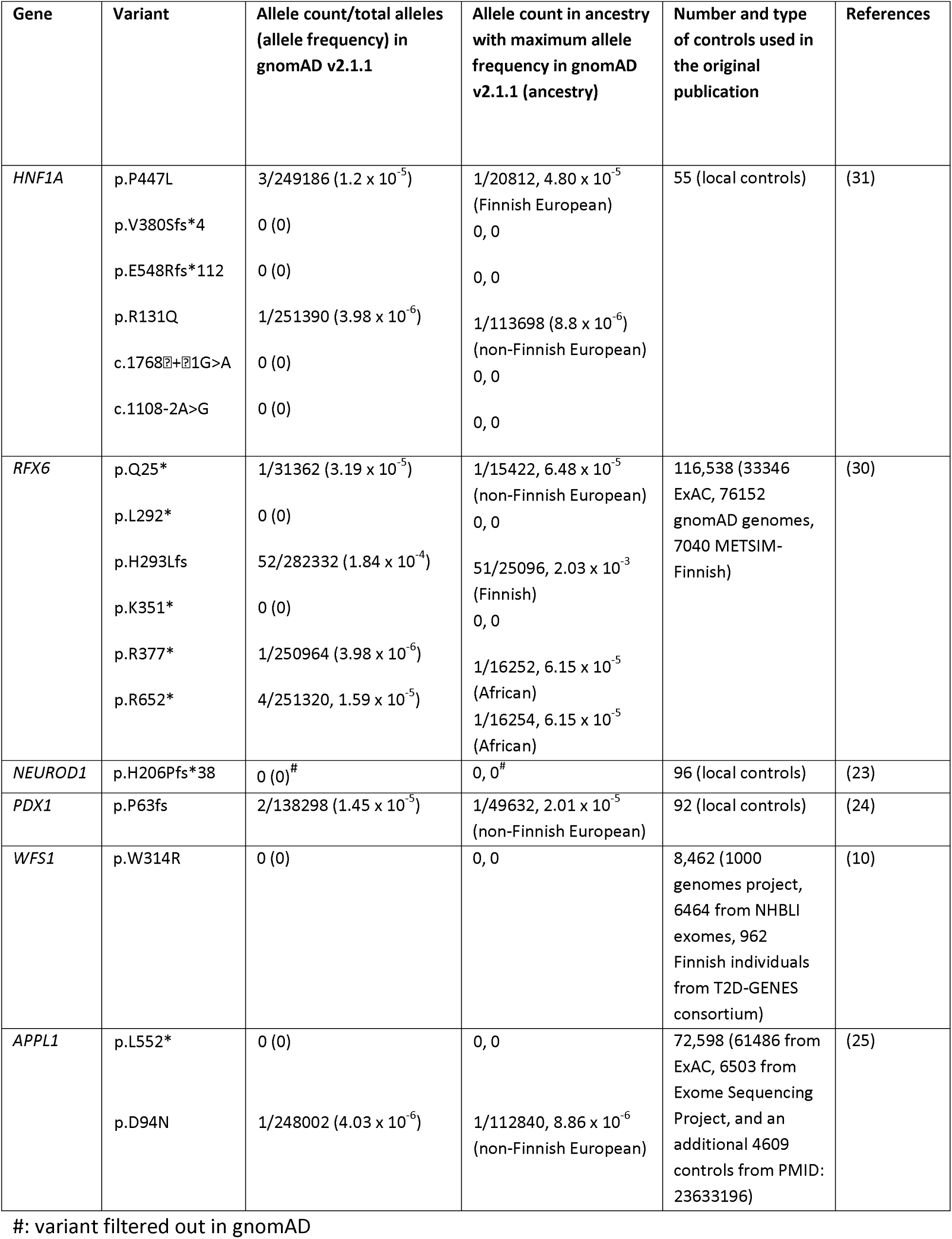
Allele frequencies of the first published variants in NEUROD1, PDX1, APPL1 and WFS1.

### All four genes show co-segregation in published pedigrees

Highly penetrant MODY-causing variants are expected to co-segregate with diabetes within families. Following ClinGen guidelines (18), we analyzed large published pedigrees with at least four co-segregations of the variant with the disease. We did not identify any additional published pedigrees for *APPL1*, *WFS1* and *PDX1* apart from the original publications. The combined logarithm of the odds (LOD) score for three families with three different *NEUROD1* variants (23,27,28) was 8.1 (Table 2). For *PDX1*, a single family showed a LOD score of 3.4 (24). *APPL1* pedigrees had a LOD score of 3.3 (26). *WFS1* demonstrated co-segregation, though with a lower LOD score of 2.1 based on one pedigree (10) (Table 2). In all these pedigrees, more than 50% of family members with diabetes had a variant (all binomial p < 0.04). However, despite its LOD score being comparable to the low-penetrance control, in the *APPL1* pedigree, more than 10% of family members aged 25 or older without diabetes carried the variant (binomial p = 0.001). This contrasted with the other genes, where non-penetrance was less frequent (p > 0.06) (Table 2).

**Table 2:**
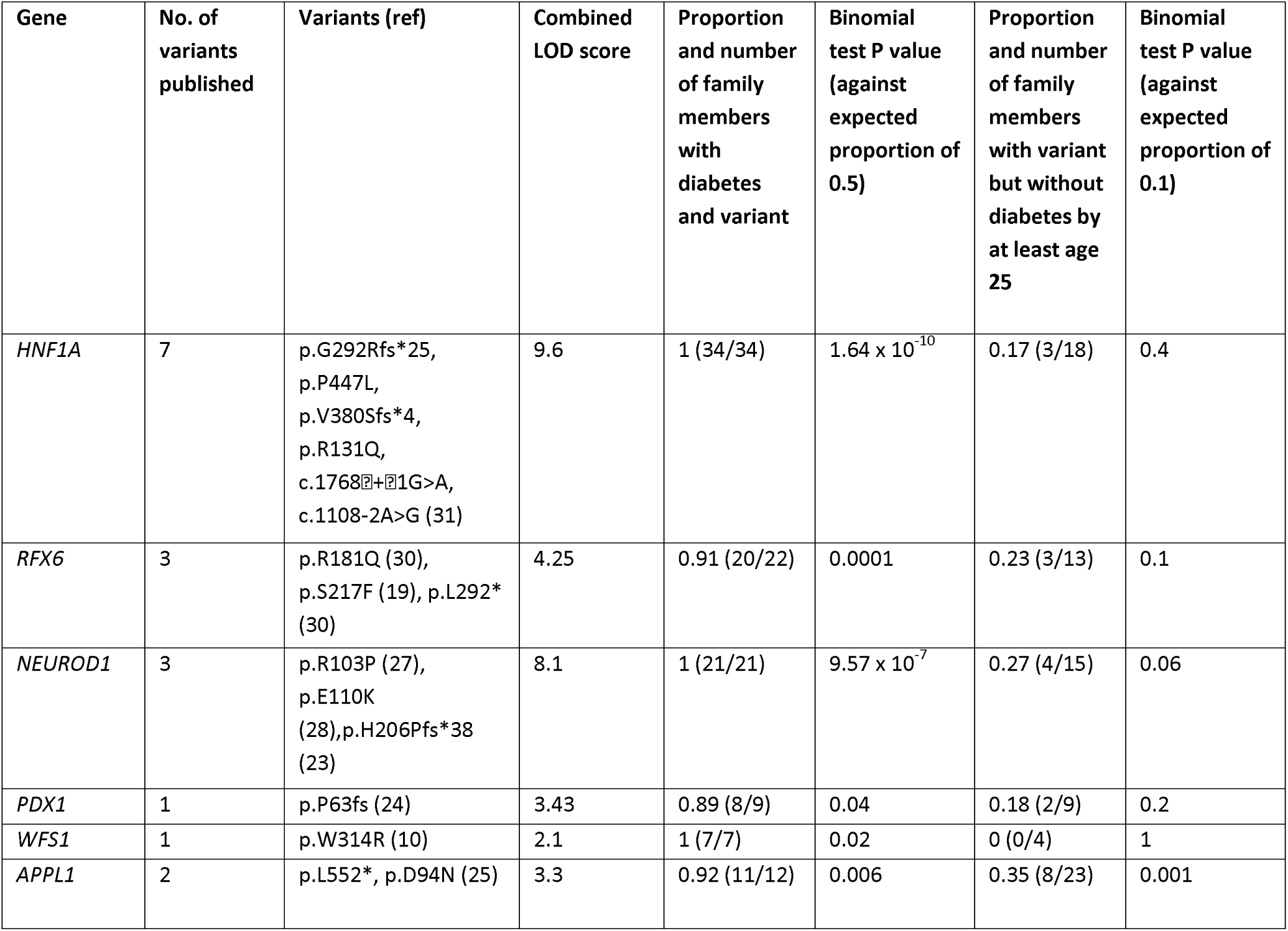
Cosegregation in published pedigrees.

### Ultra-rare PTVs and damaging missense variants are enriched in NEUROD1 and PDX1 but not in APPL1 and WFS1

Our frequency and co-segregation analysis examined specific, previously published variants and did not assess if other rare variants in these genes cause MODY. The enrichment of rare variants in a disease cohort provides strong genetic evidence of pathogenicity at the gene level. To do this, we used gene burden analyses to compare ultra-rare variants (MAF<1:10,000) in each gene between our MODY cases (n=2,471), and UK Biobank ancestry-matched controls (n=155,501). We found no enrichment of ultra-rare synonymous variants in any gene, confirming that our analysis is well calibrated, supporting our robust variant quality control process and also suggested that differences in sequencing technologies between cases and controls did not affect our results (Figure 1C, Supplementary Table 2). We observed significant enrichment of ultra-rare protein-truncating variants (PTVs) in *NEUROD1* and *PDX1* in the MODY cohort (odds ratio [OR] −21, 95% CI: 8-49, p = 3 × 10⁻⁸ and OR −30, 95% CI: 7-113, p = 3 × 10⁻⁵ respectively) (Figure 1A, Supplemental table 2). While these findings support pathogenicity, the odds ratios are closer to the low penetrance positive control *RFX6* than to the high penetrance control *HNF1A* (Figure 1A). We found similar trends for ultra-rare damaging missense variants for *NEUROD1* and *PDX1* (Figure 1A, Supplemental Table 2). In contrast, we did not find significant enrichment for ultra-rare PTVs or damaging missense variants in *APPL1* or *WFS1* (all p > 0.06, Figure 1(A and B), Supplemental Table 2).

**Figure 1:**
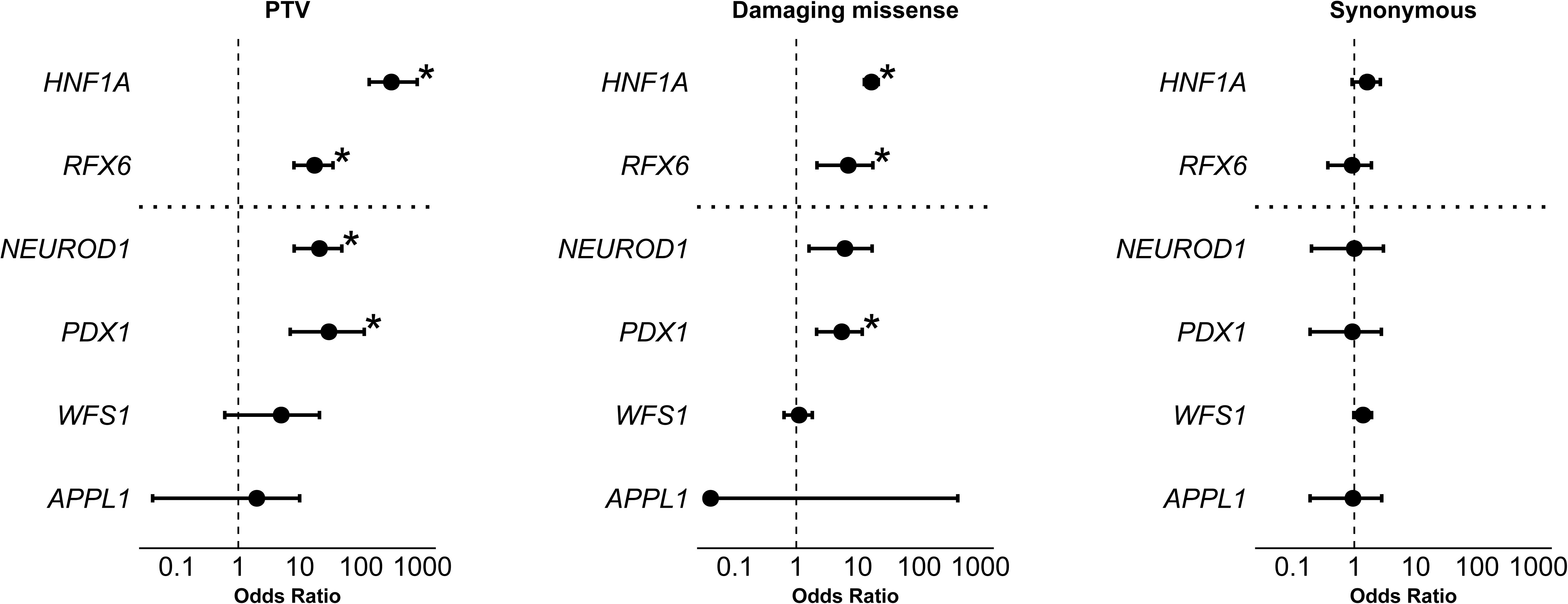
Gene-level burden analysis of ultra-rare variants in the MODY cohort. Figure shows gene-level burden analyses comparing ultra-rare variants (MAF <1×10⁻⁴) between a European-ancestry MODY cohort (n=2,471) and UK Biobank controls (n=155,501). Analyses include: (A) protein-truncating variants (B) damaging missense variants (REVEL >0.7); and (C) synonymous variants. HNF1A and RFX6 served as high- and low-penetrance positive controls, respectively. Asterisks (*) indicate significance after multiple testing corrections (p<0.002). We provide an odds ratio and a 95% confidence interval for each association.

### Multiple sensitivity analyses supported the main results that ultra-rare PTVs and damaging missense variants are enriched in NEUROD1 and PDX1 but not in APPL1 and WFS1

We conducted multiple sensitivity analyses to support our primary results. We repeated our burden tests using an alternative control set from gnomAD v3.1.1 and observed consistent findings (Supplementary Table 3). To test whether our results depended on the exact frequency threshold, we repeated the burden analyses using a stricter allele frequency threshold (MAF<1:20,000), a more lenient threshold (MAF<1:5,000), and for PTVs, we also tested not using a frequency threshold. In all cases, we observed consistent results (Supplementary Tables 4 and 5, Supplementary figure 2) that *APPL1* and *WFS1* variants are not enriched in the MODY cohort, whereas *NEUROD1* and *PDX1* were at a level similar to *RFX6*. Finally, even when limiting to variants with MAC of 1, we did not observe enrichment for *APPL1* and *WFS1* (p>0.1).

### Domain-specific analysis increases NEUROD1 and PDX1 enrichment but not for APPL1 and WFS1

We next assessed whether enrichment was restricted to specific functional regions of genes and whether including all variants, regardless of location, diluted the signal (domain information in Supplementary Table 6). We found that *WFS1* and *APPL1* showed no enrichment for PTVs or damaging missense variants whether they were within or outside functional domains (p > 0.05 for all comparisons, Figure 2). In contrast, *NEUROD1* showed stronger enrichment for ultra-rare PTVs and ultra-rare damaging missense variants within functional domains (OR: 48, 11, p < 0.0008), while variants outside these domains showed no association (p=1) despite similar length of protein within and outside the domains (174 vs. 182 amino acids). Interesting, all ultra-rare damaging missense variants in PDX1 in the MODY cohort were located within the functional domain, even though this region covered only 22% of the gene (Figure 2).

**Figure 2:**
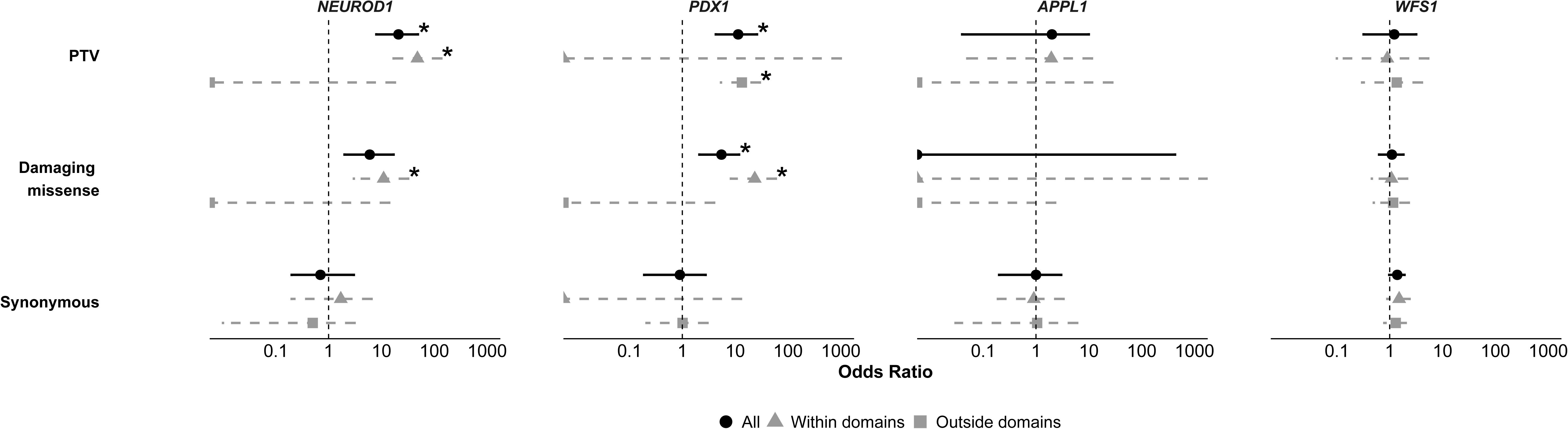
Functional domain-specific burden tests. The figure shows gene-level burden analyses comparing ultra-rare variants (MAF <1×10⁻⁴) in a European-ancestry MODY cohort (n=2,471) against UK Biobank controls (n=155,501). We show data for A) protein-truncating variants, (B) damaging missense variants (REVEL >0.7), and (C) synonymous variants. We show data for all ultra-rare variants and split them by their location within or outside the functional domains. We provide an odds ratio and a 95% confidence interval for each association. Asterisks (*) indicate significance after multiple testing corrections (p<0.002).

## Discussion

In our study using a large cohort of individuals with suspected MODY (n=2,471) and control cohorts (n=155,501), we show that ultra-rare heterozygous PTVs and damaging missense variants in *APPL1* and *WFS1* are not enriched within the MODY cohort, whereas *NEUROD1* and *PDX1* showed enrichment but at a level in line with our low-penetrance control gene *RFX6*.

The rare variants in *APPL1* are not associated with MODY. The enrichment of rare variants in a disease cohort provides strong evidence for gene-disease causality. Since the disease mechanism is expected to be loss of function, we expect to observe an enrichment of ultra-rare PTVs in the MODY cohort, similar to our positive controls, *HNF1A* and *RFX6*. However, we did not find any enrichment of ultra-rare PTVs in *APPL1*. This remains true even when focusing specifically on the functional domains, reinforcing the idea that PTVs in *APPL1* do not cause MODY. This is further supported by the prevalence of all PTVs in *APPL1* in gnomAD, which are too common to be a cause of MODY (combined allele count of 1 in 1,347 in gnomAD v4.1.0). We also did not observe an enrichment of damaging missense variants, which could potentially act through a dominant negative mechanism and might have produced a different result to PTVs. Additionally, in the *APPL1* pedigrees, 8 out of 23 individuals over the age of 25 carry the variant without having developed diabetes, five of whom are over the age of 35 (25). These data collectively suggest that the current genetic evidence does not support the causal role of *APPL1* in MODY. However, this does not imply that this gene is not biologically significant (25,31). It is also possible that rare variants may serve as a risk factor for type 2 diabetes, although the publicly available data on type 2 diabetes does not support this (P=0.60, https://t2d.hugeamp.org/gene.html?gene=APPL1).

Heterozygous variants in *WFS1* do not appear to be associated with MODY. This gene demonstrates complex genotype-phenotype relationships: recessive loss-of-function variants cause Wolfram syndrome and diabetes, C-terminal missense variants are associated with deafness, while common missense variants confer risk for type 2 diabetes. Notably, *de novo* missense variants can cause neonatal diabetes and congenital cataracts. The p.Trp314Arg variant, first reported in 2015 in a Finnish family with early-onset diabetes, was proposed to act through a loss-of-function or dominant-negative mechanism (10). Located in the first transmembrane domain (outside known functional regions), this variant shows milder effects than Wolfram syndrome-causing variants. Subsequent studies have described other heterozygous *WFS1* variants in diabetes cases, suggesting potential causality. However, our comprehensive analysis of a MODY cohort revealed no significant enrichment of ultra-rare PTVs or damaging missense variants in *WFS1*. This held true even when focusing on: (1) the functionally critical last exon, (2) known functional domains, or (3) variants with MAC=1 (comparable to p.Trp314Arg). We identified just one p.Trp314Arg carrier in our cohort (diagnosed in their 30s, BMI 22.9, no family history of diabetes, no known other pathogenic variant). These findings indicate that not all ultra-rare damaging *WFS1* variants should be considered causative for MODY. The p.Trp314Arg remains a specific case and finding from this should not be generalised to all other variants.

Evidence that variants in *NEUROD1* and *PDX1* have low-penetrance is present in the initial studies. Malecki *et al.* (23) reported all six individuals with the *NEUROD1* variant were diagnosed after 30 years of age, with the oldest in their 50s. Additionally, heterozygous parents of individuals with neonatal diabetes due to recessive PDX1 variants do not consistently have diabetes (29). This is a similar pattern to *RFX6*, an established low-penetrance cause of MODY, which had adult individuals without diabetes within the reported pedigrees (30). Consistent with this pattern of low penetrance, our burden test results for *NEUROD1* and *PDX1* are more similar to those for *RFX6* than *HNF1A*. It is interesting that enrichment was seen only in the functional domains for *NEUROD1* and for *PDX1* highlighting their critical function. These findings will need be replicated in further studies but based on our results, variants outside of the functional domain should be treated with caution.

Our study supports revising the pathogenicity classifications for these four genes. For *PDX1*, ClinGen currently assigns a genetic evidence score of 6.2/12 (based entirely on case-level data) plus 5 points for experimental evidence. Our gene burden analysis provides additional genetic evidence that would likely raise the total score above 12, warranting reclassification as “Definitive” for MODY. *NEUROD1* currently scores just 0.9 for genetic evidence and 3 for experimental evidence. Our findings could elevate its score above 7, supporting reclassification to “moderate.” While this justifies their inclusion on MODY testing panels, their low penetrance limits predictive value in unaffected individuals. *APPL1* is currently classified as “Limited” by ClinGen, while *WFS1* remains unclassified. Our gene burden results provide strong evidence against pathogenicity for MODY. We therefore recommend *APPL1* as “refuted” and restricting *WFS1* testing to specific missense variants with established evidence (10). These evidence-based revisions would optimize MODY testing strategies.

Our study has several limitations. First, our cohort was of European ancestry and we haven’t assessed other populations. While this is an important consideration, all initial reports of these four MODY-associated genes were derived from European populations, and no ancestry-specific effects have been documented for these genes to date, though future studies should explicitly evaluate these relationships in diverse ancestries. Second, although we utilized a large MODY cohort, our sample size had sufficient power only to detect associations with odds ratios ≥ 9.8 at an ultra-rare allele frequency of 0.0001. Consequently, we cannot exclude the possibility that *APPL1* or *WFS1* harbor extremely rare or low-penetrance MODY-causing variants. However, the relatively high population frequency of damaging variants in these genes suggests they are more likely to act as type 2 diabetes risk factors rather than monogenic causes of MODY.

In conclusion, we provide genetic evidence that *NEUROD1* and *PDX1* cause low penetrance MODY whereas variants in *APPL1* and *WFS1* do not cause MODY and should not be included in MODY genetic analysis. Our study highlights the importance of re-evaluating gene-disease relationships in light of available evidence, particularly the benefit of gene level rare variant analysis to support the genetic evidence of pathogenicity.

## Supporting information

Supplemental methods and results

## Data Availability

All data produced in the present work are contained in the manuscript

## Acknowledgments

This research has been conducted using the UK Biobank Resource. This work was conducted under UK Biobank project number 9072. KAP is funded by the Wellcome Trust (219606/Z/19/Z). T.W.L is supported by the Academy of Medical Sciences/the Wellcome Trust/the Government Department of Science Innovation and Technology/the British Heart Foundation/Diabetes UK Springboard Award [SBF009\1135]. ATH is supported by Wellcome Trust Senior Investigator award (WT098395/Z/12/Z). The work is supported by the National Institute for Health Research (NIHR) Exeter Biomedical Research Centre, Exeter, UK. And NIHR Exeter Clinical Research Facilities. The Wellcome Trust, MRC and NIHR had no role in the design and conduct of the study; collection, management, analysis, and interpretation of the data; preparation, review, or approval of the manuscript; and decision to submit the manuscript for publication. The views expressed are those of the author(s) and not necessarily those of the Wellcome Trust, Department of Health, NHS or NIHR. For the purpose of open access, the author has applied a CC BY public copyright licence to any Author Accepted Manuscript version arising from this submission.

## Funding

European Foundation for the Study of Diabetes.

## Duality of Interest

The authors have no conflicts of interest to declare.

## Author Contributions

A.S: Formal analysis, Writing - Original Draft, Writing - Review & Editing, Visualization. M.N.Wa: Methodology, Formal analysis, Writing - Review & Editing. A.T.H: Resources, Supervision. M.N.We: Conceptualization, Methodology, Writing - Review & Editing, Funding acquisition. K.C: Resources, Data Curation, Writing - Review & Editing. T.W.L: Conceptualization, Methodology, Formal analysis, Writing - Original Draft, Writing - Review & Editing, Funding acquisition. K.A.P: Conceptualization, Methodology, Formal analysis, Resources, Writing - Original Draft, Writing - Review & Editing, Supervision, Funding acquisition. K.A.P is the guarantor of this work and, as such, had full access to all the data in the study and takes responsibility for the integrity of the data and the accuracy of the data analysis.

## Notes

### Competing Interest Statement

The authors have declared no competing interest.

### Funding Statement

This study was funded by the Wellcome Trust and the European Foundation for the Study of Diabetes.

### Author Declarations

The North Wales Ethics committee gave ethical approval for this work (no. 17/WA/0327).

