## Supplemental methods and results for "*NEUROD1* and *PDX1* are low penetrance causes of MODY while rare variants in *APPL1* and *WFS1* are not associated with MODY"

**Ancestry**

We determined ancestry in the MODY cohort using an implementation of the LASER method for targeted panel data (1).

**Quality Control**

**MODY cohort**

We implemented a rigorous quality control (QC) process at the sample, variant, and genotype levels to ensure the integrity of sequencing data obtained from different sequencing technologies. Given the potential discrepancies that arise when comparing sequencing data from multiple sources, applying stringent QC measures was essential to minimize errors and enhance reliability. We removed six samples due to higher missingness of >2%. We performed multiple QC steps at variant level that included removing variants with (A) an FS (Phred-scaled p-value using Fisher's exact test for strand bias) greater than 60, (B) a QD (Variant Confidence/Quality by Depth) lower than 2, (C) a ReadPosRankSum (Z-score from the Wilcoxon rank sum test of Alt vs. Ref read position bias) lower than -8, and (D) an MQRanksum (Z-score from the Wilcoxon rank sum test of Alt vs. Ref read mapping qualities) lower than -12.5. We also removed variants with a (E) mapping quality lower than 40, a (F) read depth below 20, or (G) a genotype quality below 20. Additionally, we filtered out variants with allelic imbalance, if a binomial test of detecting allelic imbalance yielded a p-value less than 0.001. Finally, we removed the genotypes with a missingness rate exceeding 2%.

After implementing all QC measures, we retained 660 high-quality variants, ensuring a robust dataset for downstream analysis.

**UK Biobank**

We performed the same QC in the UK Biobank with minor changes to reflect it had genome sequencing rather targeted gene panel data. The changes included lowering the read depth to 15 for excluding the variants. We also removed variants which had poor AA score (< 0.5). The AAscore is a quality metric that estimates the probability of a variant being a true positive, generated by Graphtyper (2)Finally, we removed variants which are flagged as being located in low-complexity regions by gnomAD to remove potentially false positive variants. Any variant that failed QC in either MODY or UK biobank were removed from both cohorts for the analysis.

**GnomAD**

Gnomad has already performed sample and variant QC. In addition to this, we excluded variants if they were located in regions with low coverage (≤10× in ≤80% of samples). We also excluded variants that were filtered by gnomAD and also removed variants if they were flagged as being located in a low complexity region in gnomAD. Any variant that failed QC in either the MODY cohort or gnomAD were removed from both cohorts for the analysis.

**Supplemental Results**

Supplemental table 1: Characteristics of MODY cohort

Median (IQR) for continuous variable and n (%) for categorical data

| **Characteristics** | **MODY cohort** |
| --- | --- |
| N | 2471 |
| Age of diagnosis of diabetes, years | 23 (16-31) |
| Female Sex | 59.4% |
| Age at recruitment, years | 32 (22-42) |
| BMI, kg/m^2^ | 26 (22.7-30.2) |
| Parents with diabetes | 67.5% |
| HbA1c, % | 7.8 (6.7-10) |
| On Insulin treatment | 47.6% |
| European ancestry (genetically determined) | 100% |

**Supplemental table 2: Gene burden test for variants with MAF < 0.0001 with UK biobank as controls**

| **Variant type** | **Gene** | **Allele count  in MODY  cohort** | **Allele count  in population  cohort  (UK biobank)** | **Odds ratio (95% CI)** | **P Value** |
| --- | --- | --- | --- | --- | --- |
| Rare PTVs | *HNF1A* | 36 | 7 | 313 (136-824) | 1.31 x 10^-57^ |
| (MAF < 0.0001) | *RFX6* | 11 | 38 | 17 (8-35) | 3.55 x 10^-10^ |
|  | *NEUROD1* | 8 | 23 | 21 (8-49) | 2.80 x 10^-8^ |
|  | *PDX1* | 4 | 8 | 30 (7-113) | 3.13 x 10^-5^ |
|  | *WFS1* | 2 | 23 | 5 (0.6-21) | 0.06 |
|  | *APPL1* | 1 | 47 | 2 (0.04-10) | 0.5 |
| Rare damaging missense variants | *HNF1A* | 78 | 281 | 17 (13-22) | 1.14 x 10^-61^ |
| (MAF < 0.0001) | *RFX6* | 5 | 43 | 7 (2-18) | 0.001 |
|  | *NEUROD1* | 4 | 39 | 6 (2-17) | 0.005 |
|  | *PDX1* | 7 | 77 | 5 (2-12) | 0.0005 |
|  | *WFS1* | 16 | 868 | 1 (0.6-2) | 0.6 |
|  | *APPL1* | 0 | 2 | 0 (0-429) | 1 |
| All PTVs | *HNF1A* | 36 | 7 | 313 (136-824) | 1.31 x 10^-57^ |
|  | *RFX6* | 11 | 38 | 17 (8-35) | 3.55 x 10^-10^ |
|  | *NEUROD1* | 8 | 23 | 21 (8-49) | 2.80 x 10^-8^ |
|  | *PDX1* | 4 | 8 | 30 (7-113) | 3.13 x 10^-5^ |
|  | *WFS1* | 16 | 845 | 1 (0.6-2) | 0.6 |
|  | *APPL1* | 6 | 255 | 2 (0.7-4) | 0.1 |
| Rare synonymous variants | *HNF1A* | 16 | 598 | 2 (1-3) | 0.08 |
| (MAF < 0.0001) | *RFX6* | 7 | 462 | 1 (0.4-2) | 1.00 |
|  | *NEUROD1* | 3 | 181 | 1 (0.2-3) | 1.00 |
|  | *PDX1* | 3 | 195 | 1 (0.2-3) | 1.00 |
|  | *WFS1* | 39 | 1695 | 1 (0.98-2) | 0.05 |
|  | *APPL1* | 3 | 252 | 1 (0.2-3) | 1.00 |

**Supplemental table 3: Gene burden test for variants with MAF < 0.0001 with gnomAD v3 as controls**

| **Variant type** | **Gene** | **Allele count  in MODY  cohort** | **Allele count  in population  cohort  (gnomAD)** | **Odds ratio (95% CI)** | **P Value** |
| --- | --- | --- | --- | --- | --- |
| Rare PTVs | *HNF1A* | 37 | 3 | 163 (52-815) | 1.92 x 10^-39^ |
| (MAF < 0.0001) | *RFX6* | 9 | 9 | 13 (5-38) | 1.14 x 10^-6^ |
|  | *NEUROD1* | 8 | 6 | 18 (5-62) | 1.22x10^-6^ |
|  | *PDX1* | 4 | 4 | 13 (2-71) | 0.001 |
|  | *WFS1* | 2 | 7 | 4 (0.4-20) | 0.1 |
|  | *APPL1* | 1 | 5 | 3 (0.1-31) | 0.3 |
| Rare damaging missense variants | *HNF1A* | 78 | 61 | 17 (12-24) | 3.99 x 10^-52^ |
| (MAF < 0.0001) | *RFX6* | 5 | 1 | 66 (7-3067) | 0.00001 |
|  | *NEUROD1* | 4 | 9 | 6 (1-21) | 0.01 |
|  | *PDX1* | 7 | 9 | 10 (3-31) | 0.0001 |
|  | *WFS1* | 19 | 182 | 1 (1-2) | 0.2 |
|  | *APPL1* | 0 | 0 | 0 (0-Inf) | 1 |
| All PTVs | *HNF1A* | 37 | 3 | 163 (52-815) | 1.92 x 10^-39^ |
|  | *RFX6* | 9 | 9 | 13 (5-38) | 1.14 x 10^-6^ |
|  | *NEUROD1* | 8 | 6 | 18 (5-62) | 1.22 x 10^-6^ |
|  | *PDX1* | 4 | 4 | 13 (2-71) | 0.001 |
|  | *WFS1* | 3 | 17 | 2 (0.4-8) | 0.2 |
|  | *APPL1* | 6 | 54 | 2 (1-5) | 0.1 |
| Rare synonymous variants | *HNF1A* | 15 | 132 | 2 (1-3) | 0.14 |
| (MAF < 0.0001) | *RFX6* | 9 | 102 | 1 (1-2) | 0.58 |
|  | *NEUROD1* | 3 | 59 | 1 (0.1-2) | 0.80 |
|  | *PDX1* | 3 | 49 | 1 (0.2-3) | 1.00 |
|  | *WFS1* | 35 | 348 | 1 (1-2) | 0.13 |
|  | *APPL1* | 2 | 56 | 1 (0.1-2) | 0.77 |

**Supplemental table 4: Sensitivity analysis burden test (MAF<0.00005) in MODY cohort (n = 2,471) and UK Biobank (n = 155,501)**

| **Variant type** | **Gene** | **Allele count  in MODY  cohort** | **Allele count  in population  cohort  (UK biobank)** | **Odds ratio (95% CI)** | **P Value** |
| --- | --- | --- | --- | --- | --- |
| Rare PTVs | *HNF1A* | 36 | 7 | 313 (136-824) | 1.31 x 10^-57^ |
| (MAF < 0.00005) | *RFX6* | 11 | 38 | 17 (8-35) | 3.55 x 10^-10^ |
|  | *NEUROD1* | 8 | 23 | 21 (8-49) | 2.80 x 10^-8^ |
|  | *PDX1* | 4 | 8 | 30 (7-113) | 3.13 x 10^-5^ |
|  | *WFS1* | 2 | 23 | 5 (0.6-21.2) | 0.06 |
|  | *APPL1* | 1 | 47 | 2 (0.04-10) | 0.5 |
| Rare damaging missense variants | *HNF1A* | 77 | 240 | 19 (15-25) | 7.24 x 10^-65^ |
| (MAF < 0.00005) | *RFX6* | 5 | 43 | 7 (2-18) | 0.001 |
|  | *NEUROD1* | 4 | 39 | 6 (2-17) | 0.005 |
|  | *PDX1* | 7 | 32 | 13 (5-31) | 2.96 x 10^-6^ |
|  | *WFS1* | 12 | 533 | 1 (1-2) | 12 |
|  | *APPL1* | 0 | 2 | 0 (0-429) | 1 |
| Rare synonymous variants | *HNF1A* | 11 | 395 | 2 (1-3) | 0.11 |
| (MAF < 0.00005) | *RFX6* | 5 | 268 | 1 (0.4-3) | 0.64 |
|  | *NEUROD1* | 2 | 122 | 1 (0.1-4) | 1.00 |
|  | *PDX1* | 3 | 178 | 1 (0.2-3) | 0.77 |
|  | *WFS1* | 27 | 964 | 2 (1-2) | 0.01 |
|  | *APPL1* | 1 | 209 | 0 (0-2) | 0.53 |

**Supplemental table 5: Sensitivity analysis burden test (MAF<0.0002) in MODY cohort (n = 2,471) and UK Biobank (n = 155,501)**

| **Variant type** | **Gene** | **Allele count  in MODY  cohort** | **Allele count  in population  cohort  (UK biobank)** | **Odds ratio (95% CI)** | **P Value** |
| --- | --- | --- | --- | --- | --- |
| Rare PTVs | *HNF1A* | 36 | 7 | 313 (136-824) | 1.31 x 10^-57^ |
| (MAF < 0.0002) | *RFX6* | 11 | 38 | 17 (8-35) | 3.55 x 10^-10^ |
|  | *NEUROD1* | 8 | 23 | 21 (8-49) | 2.80 x 10^-8^ |
|  | *PDX1* | 4 | 8 | 30 (7-113) | 3.13 x 10^-5^ |
|  | *WFS1* | 2 | 23 | 5 (0.6-21.2) | 0.06 |
|  | *APPL1* | 1 | 47 | 2 (0.04-10) | 0.5 |
| Rare damaging missense variants | *HNF1A* | 81 | 372 | 13 (10-17) | 6.56 x 10^-57^ |
| (MAF < 0.0002) | *RFX6* | 5 | 43 | 7 (2-18) | 0.001 |
|  | *NEUROD1* | 4 | 39 | 6 (2-17) | 0.005 |
|  | *PDX1* | 7 | 77 | 6 (2-12) | 0.0005 |
|  | *WFS1* | 34 | 1296 | 2 (1-2) | 0.01 |
|  | *APPL1* | 0 | 2 | 0 (0-429) | 1 |
| Rare synonymous variants | *HNF1A* | 22 | 882 | 2 (1-2) | 0.06 |
| (MAF < 0.0002) | *RFX6* | 11 | 678 | 1 (0.5-2) | 1.00 |
|  | *NEUROD1* | 4 | 218 | 1 (0.3-3) | 0.79 |
|  | *PDX1* | 3 | 240 | 1 (0.2-2) | 1.00 |
|  | *WFS1* | 46 | 2233 | 1 (1-2) | 0.16 |
|  | *APPL1* | 4 | 393 | 1 (0.2-2) | 1.00 |

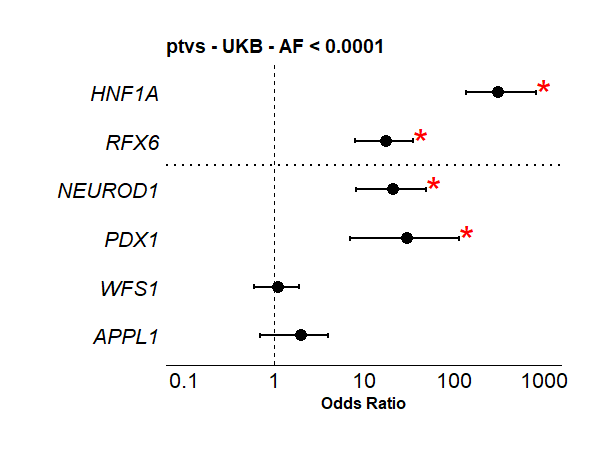

**Supplemental figure 1: Gene burden test for all protein truncating variants in MODY cohort (n = 2,471) and UK Biobank (n = 155,501).** Analysis includes all protein-truncating variants, with no allele frequency threshold. *HNF1A* and *RFX6* served as high- and low-penetrance positive controls, respectively. Asterisks (*) indicate significance after multiple testing corrections (p<0.002). We provide an odds ratio and a 95% confidence interval for each association.

**Supplemental table 6: Functional domains used in domain specific burden test analyses**

| **Gene** | **Uniprot ID** | **Domains:**  **Interpro* representative domains + Pfam domain** | **Domain sizes combined/gene size** |
| --- | --- | --- | --- |
| *APPL1* | Q9UKG1 | BAR domain of APPL family: 7-247,  Adaptor protein containing PH domain: 252-376  Phosphotyrosine-binding domain: 497-636 | 503/709 (71%) |
| *WFS1* | O76024 | Wolframin Sel1-like repeat: 99-133, 137-175,  Wolframin EF-hand domain: 177-255,  Wolframin cysteine-rich domain: 666-769  Wolframin C-terminal OB-fold domain: 770-889 | 372/890 (42%) |
| *NEUROD1* | Q13562 | Basic helix-loop-helix: 75-160  Neuronal helix-loop-helix transcription factor: 160-284 | 209/356 (59%) |
| *PDX1* | P52945 | Homeodomain: 146-208 | 62/283 (22%) |

*InterPro 104.0. Accessed on 5/3/2025

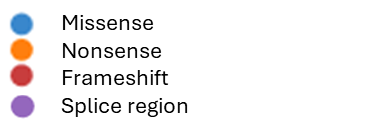

***NEUROD1***

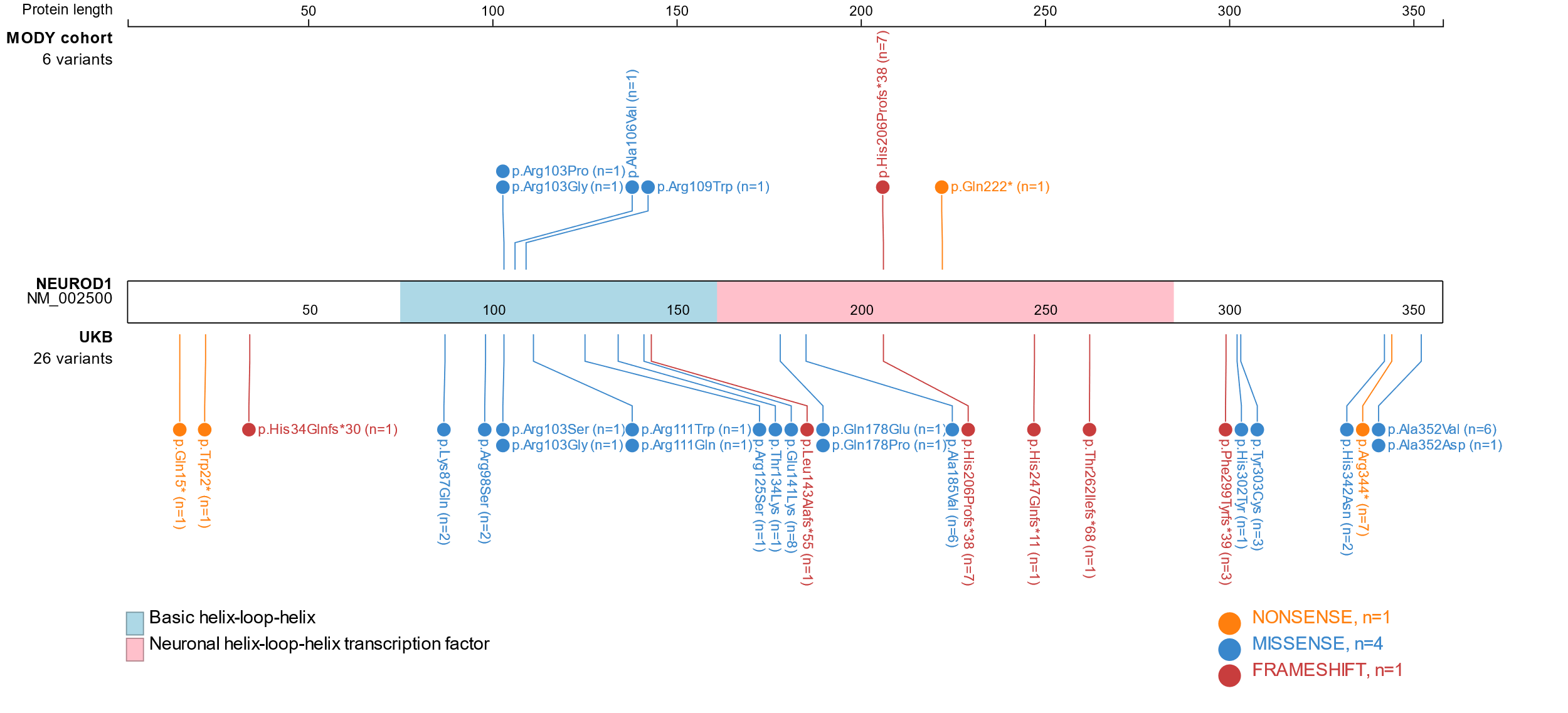

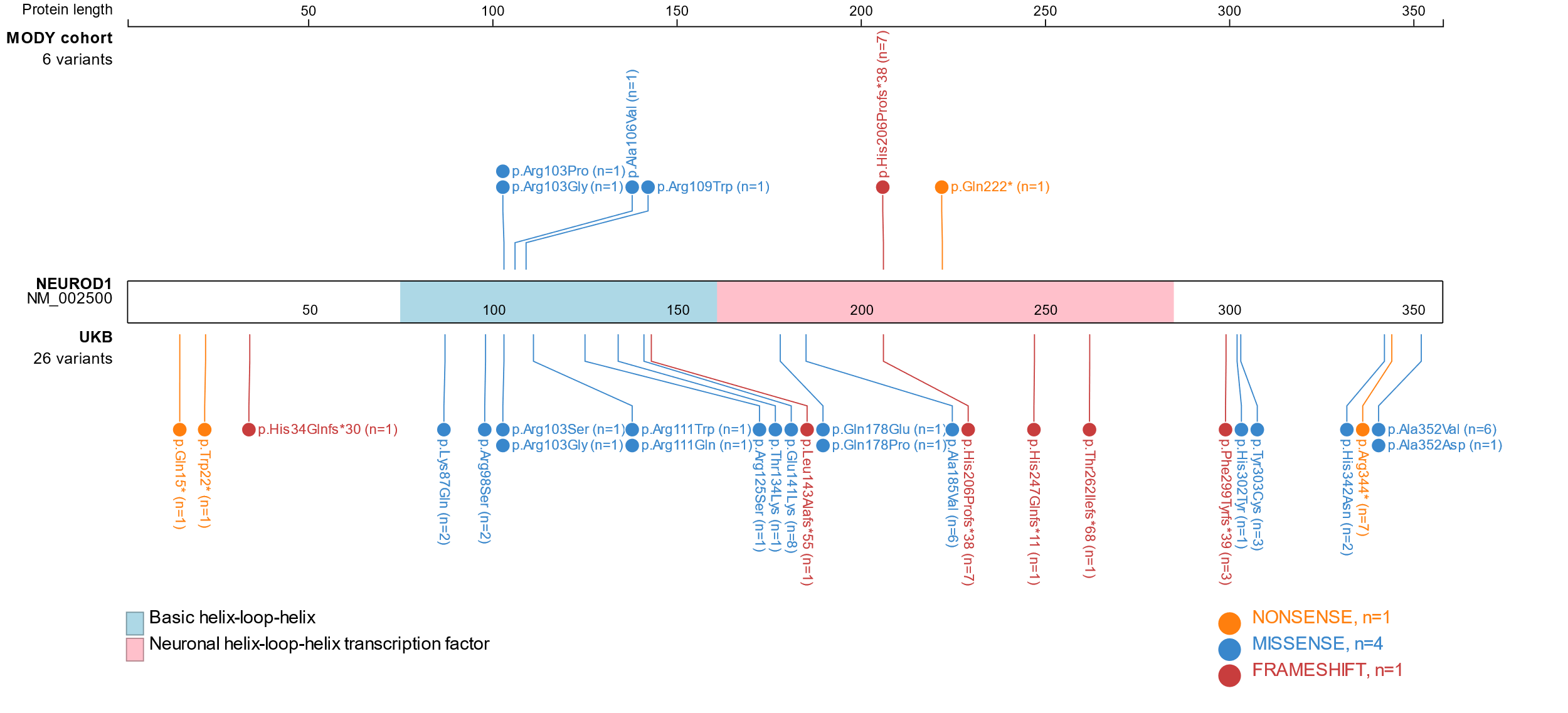

***PDX1***

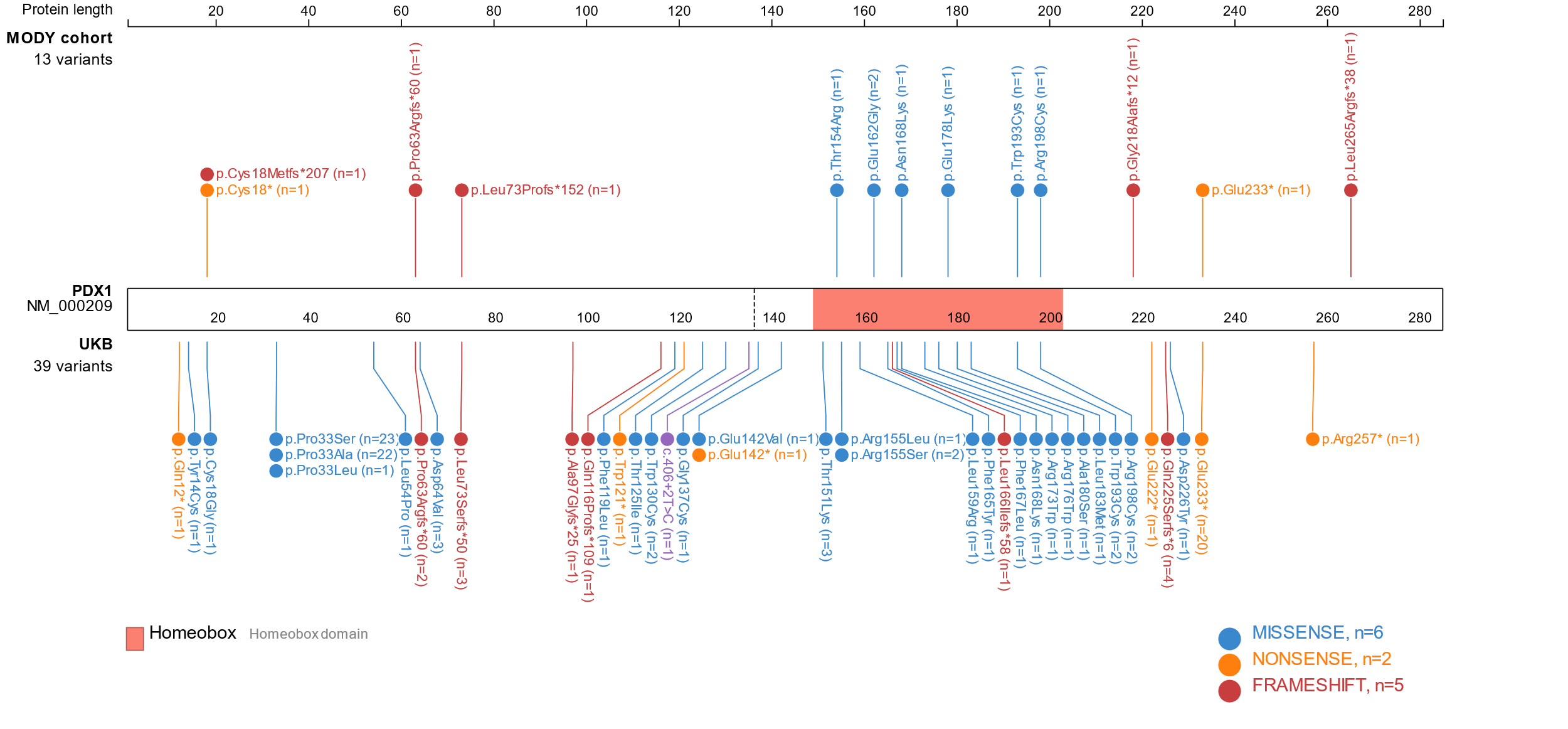

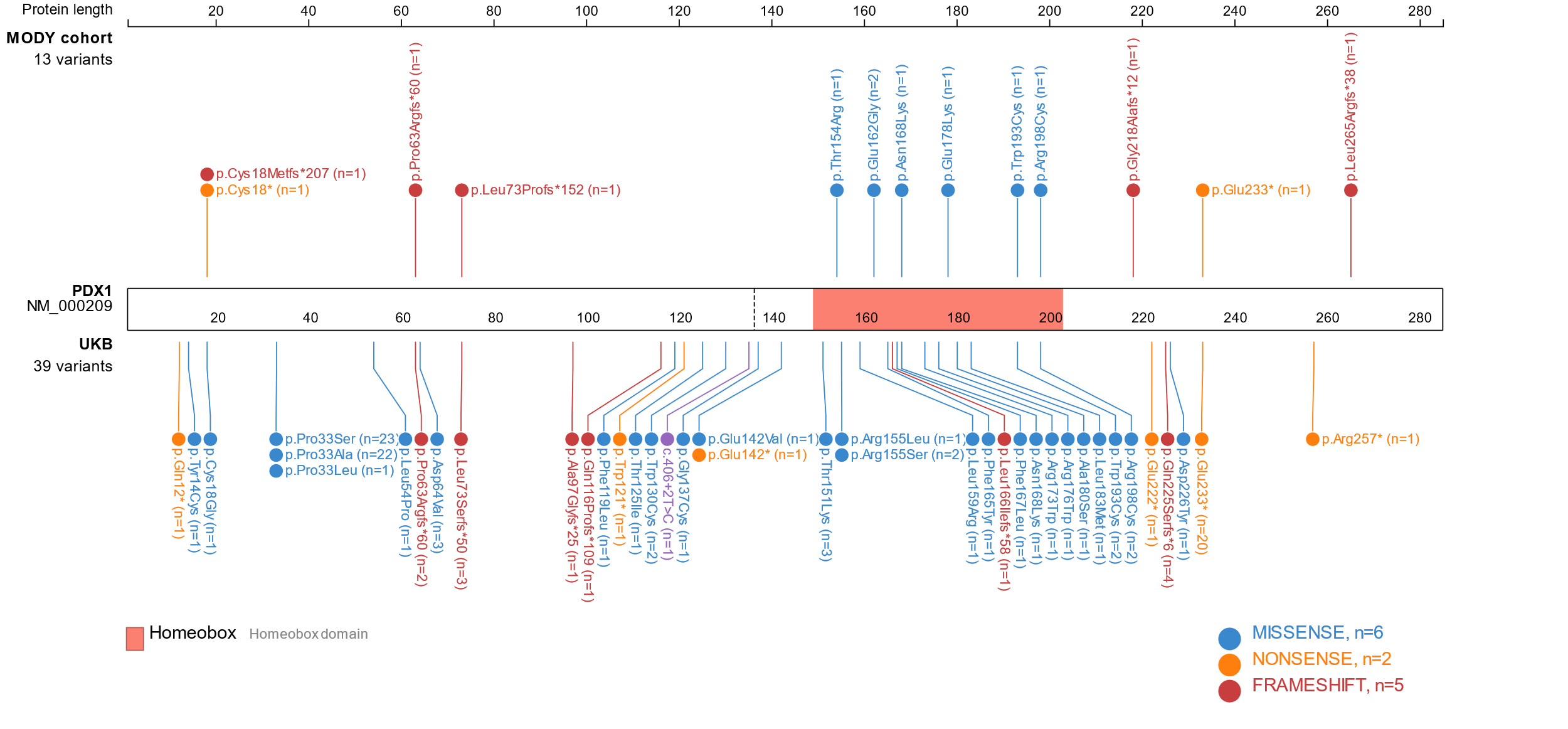

***WFS1***

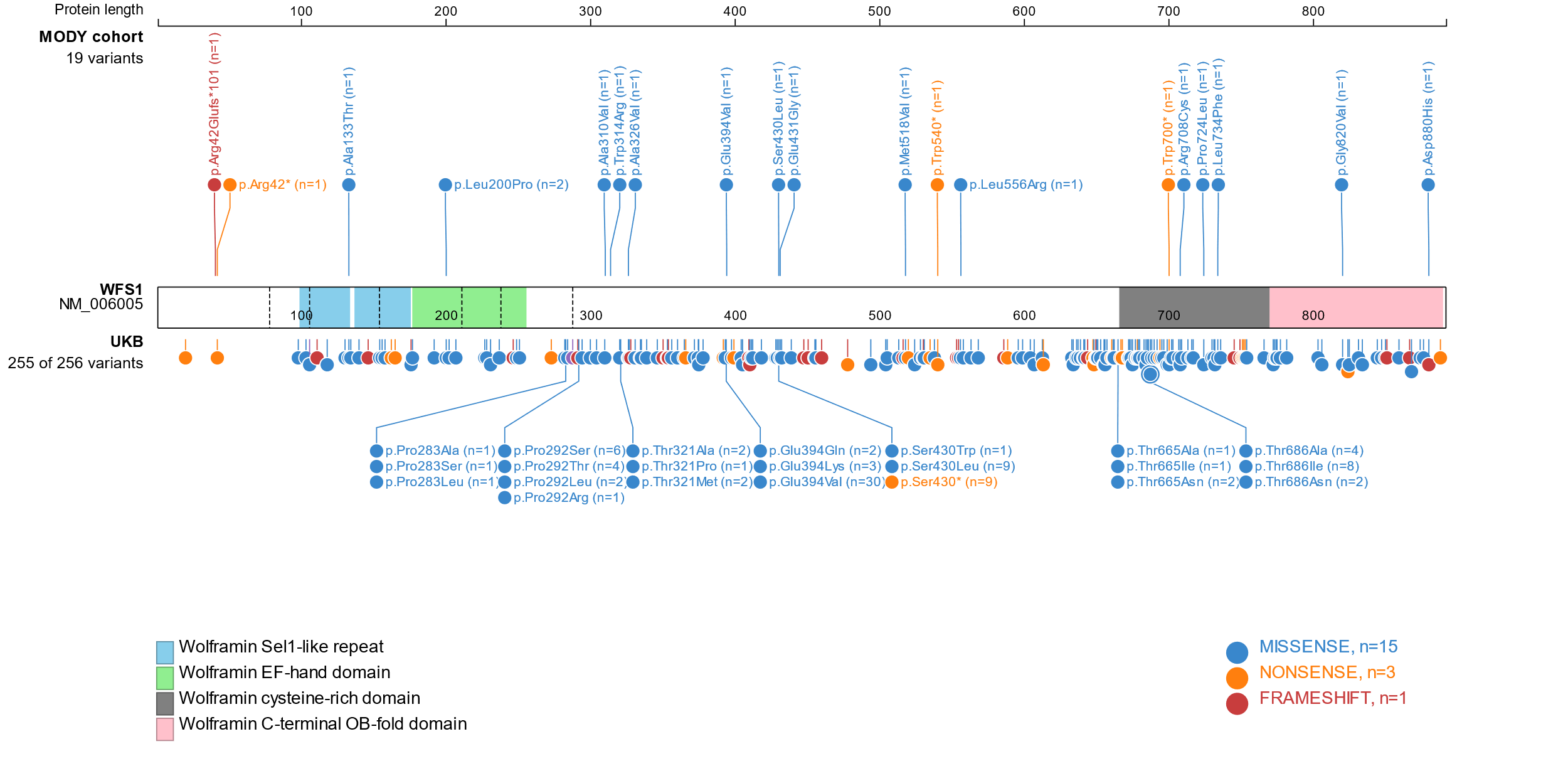

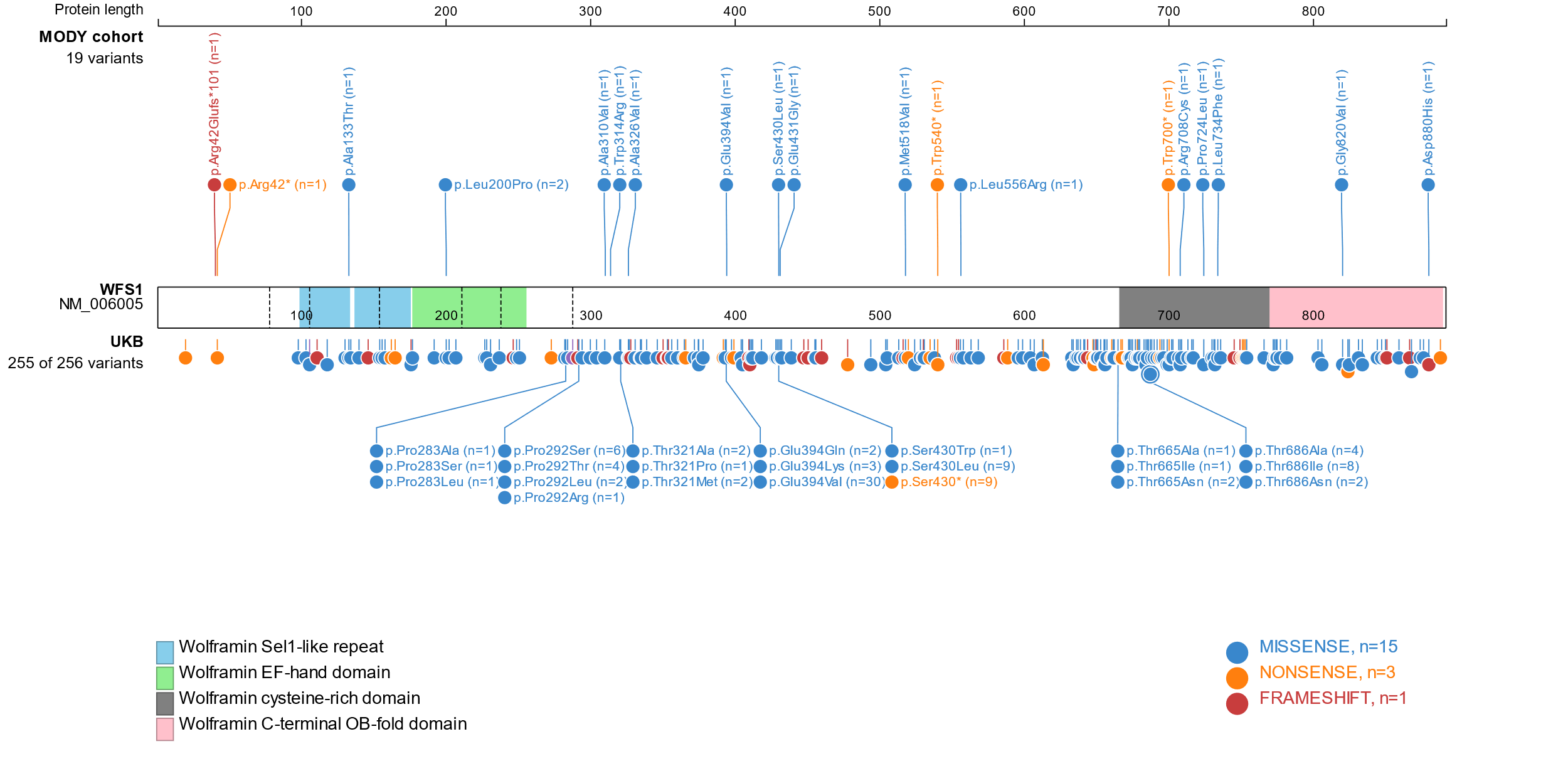

***APPL1***

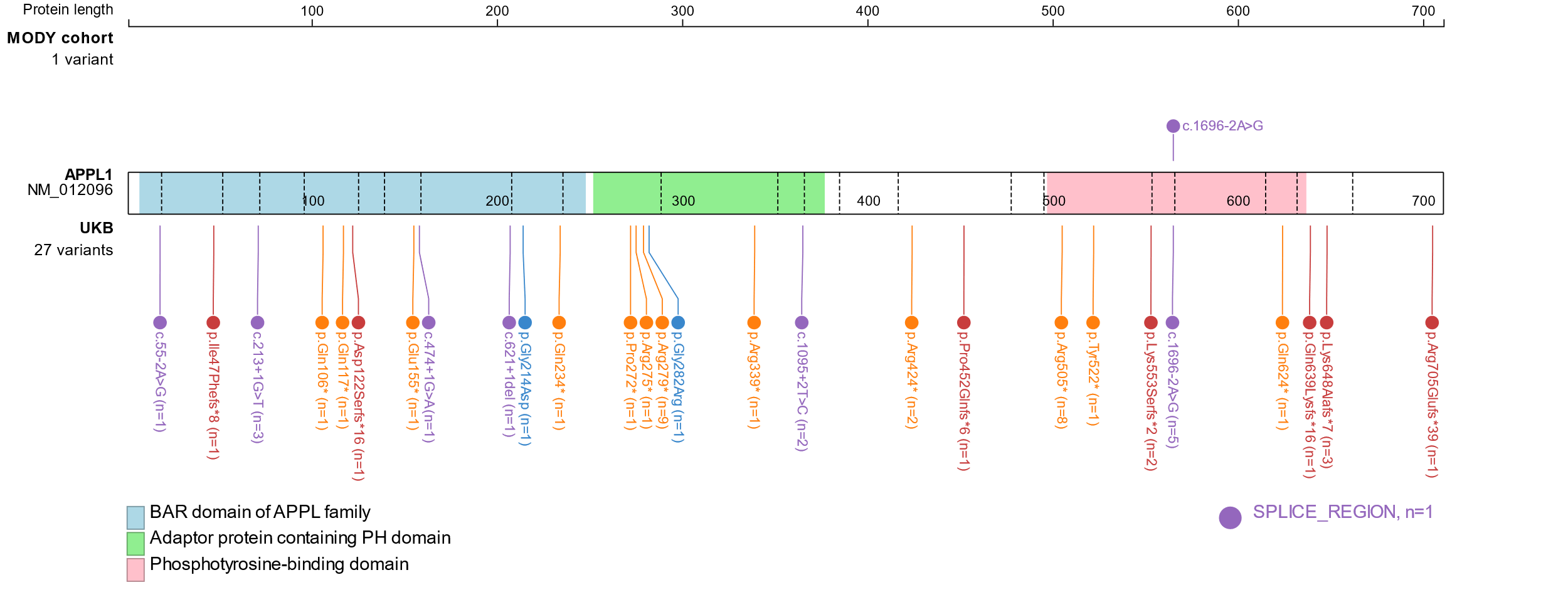

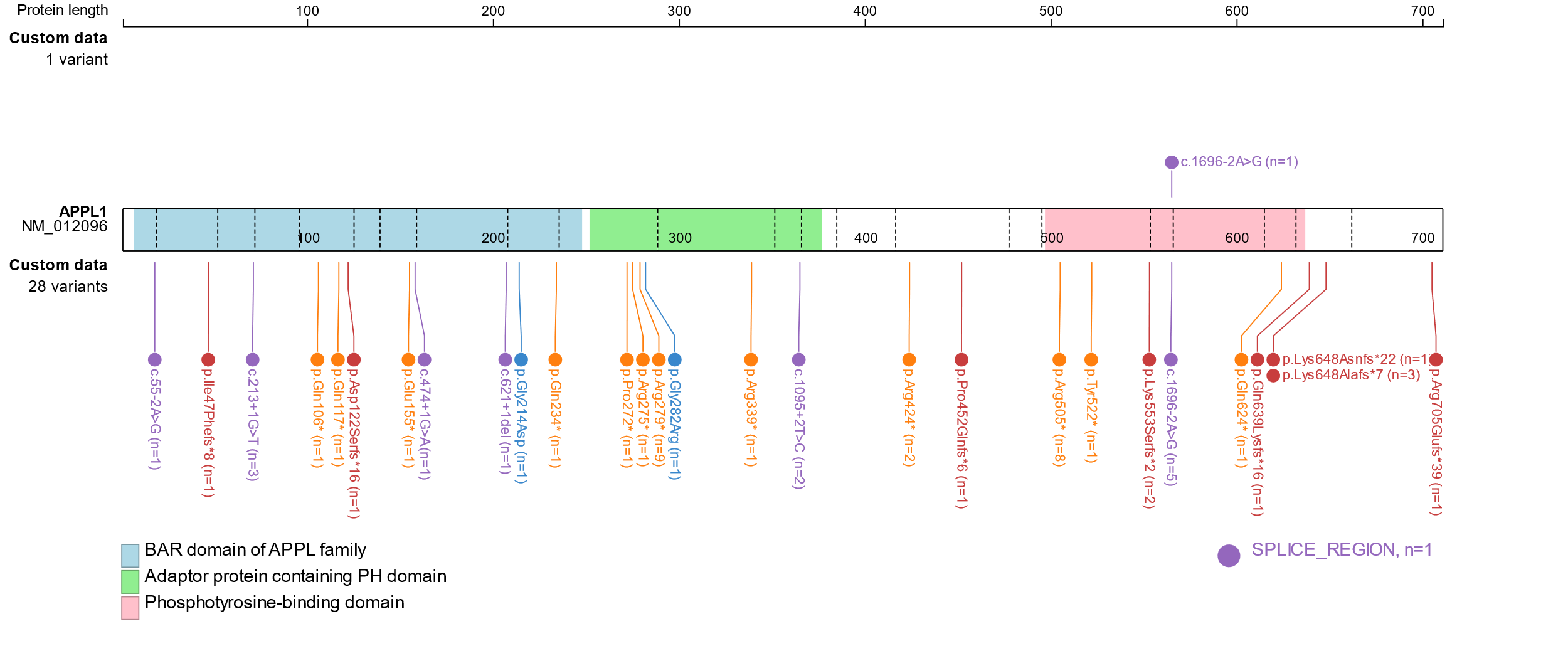

**Supplementary figure 2: Distribution of rare variants (MAF < 0.0001) in *NEUROD1*, *PDX1*, *APPL1* and *WFS1*.** This includes protein truncating variants split into nonsense, frameshift and splice region variants, and missense variants with a REVEL score > 0.7. The functional domains within each gene have been indicated. Variants identified in the MODY cohort are shown above the gene, while those from the UK Biobank are displayed below. Figures were generated using ProteinPaint (<https://proteinpaint.stjude.org/>).

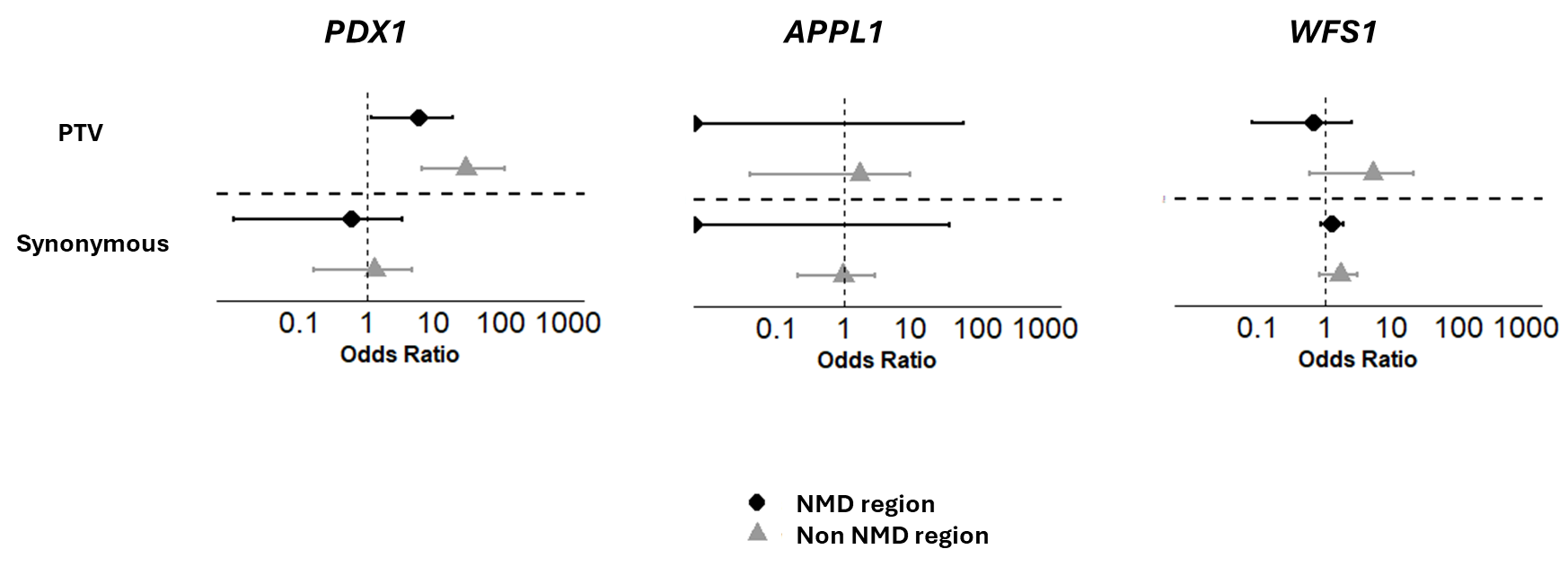

**Supplementary figure 3: Gene burden tests for rare (MAF < 0.0001) protein truncating variants (PTV) and synonymous variants within and outside the NMD regions in *PDX1*, *APPL1* and *WFS1*.** This compares the MODY cohort (n = 2,471) and UK Biobank (n = 155,501). NMD refers to nonsense-mediated decay. The non-NMD region analysis included only PTVs in the last exon and the last 50bp of the penultimate exon of our genes of interest, while the NMD region analysis included PTVs in the rest of the gene. Synonymous variants were tested as a sensitivity analysis. *NEUROD1* was excluded because it is a single exon gene with all positions predicted to escape NMD.
